# Intramyocardial cell-based therapy during bidirectional cavopulmonary anastomosis for hypoplastic left heart syndrome: The ELPIS phase I trial

**DOI:** 10.1101/2022.08.04.22278321

**Authors:** Sunjay Kaushal, Jessica R Hoffman, Riley M Boyd, Joshua M Hare, Kevin N. Ramdas, Nicholas Pietris, Shelby Kutty, James S Tweddell, S. Adil Husain, Shaji C. Menon, Linda M. Lambert, David A Danford, Seth J Kligerman, Narutoshi Hibino, Laxminarayana Korutla, Prashanth Vallabhajosyula, Michael J. Campbell, Aisha Khan, Keyvan Yousefi, Danial Mehranfard, Lisa McClain-Moss, Anthony A. Oliva, Michael E Davis

## Abstract

**Background:** Hypoplastic left heart syndrome (HLHS) survival relies on surgical reconstruction for the right ventricle (RV) to provide systemic circulation. This leads to substantially increased loads on the RV, wall stress, maladaptive remodeling and dysfunction, which in turn can increase risk of death or transplantation.

**Objectives:** We conducted a phase I multicenter trial to assess safety and feasibility of intra-operative MSC injection in HLHS patients to boost RV performance in the systemic position.

**Methods:** Allogeneic MSCs were directly administered by intramyocardial injections during the second stage palliative operation. The primary endpoint was safety.

**Results:** Ten patients received intramyocardial injections of allogeneic MSCs (Lomecel-B). No patients experienced major adverse cardiac events (MACE). All subjects were alive and transplant-free at 1 year following, and experienced growth comparable to healthy control historical data. Cardiac magnetic resonance imaging (CMR) revealed improving tricuspid regurgitant fraction (Baseline: 0.45±0.19; 6 mo.: 0.32±0.06; 12 mo.: 0.06±0.09), while global longitudinal strain (Baseline: -24.39±6.99; 6 mo.: -20.55±3.05, *p* > 0.05 vs baseline; 12 mo.: - 23.88±4.6, *p*>0.05 vs baseline) and RV ejection fraction (EF; baseline: 62.62±5.99; 6 mo.: 53.69±9.56; 12 mo.: 52.31±5.63: *p=NS* for change over time) were unchanged. Computational modeling identified 167 derived RNAs specific to circulating exosomes originating from transplanted MSCs corresponding to RVEF changes and identifying potential mechanistic underpinnings.

**Conclusions:** Intramyocardial MSCs appear safe in HLHS patients, and may favorably affect RV performance. Circulating exosomes of transplanted MSC-specific provide novel insight into bioactivity. Conduct of a controlled phase trial is warranted and is underway.

**Condensed Abstract:** The ELPIS phase I trial was designed to assess safety and feasibility of intramyocardial injection of allogeneic MSCs into the RV during second stage palliation of HLHS. There were no incidences of major adverse cardiac events (MACE) or other safety concerns, and there was a 100% transplant-free survival at 1-year follow-up, supporting the safety and feasibility of this approach. The ELPIS results are important for advancing MSC therapy for all ages and congenital heart conditions, and warrant further investigation in a controlled Phase II trial powered for efficacy.

## INTRODUCTION

Hypoplastic left heart syndrome (HLHS) is one of the most severe and complex congenital heart defects that inevitably leads to death without intervention (1). With an incidence of approximately 2-3.5 per 10,0000 live births (2–4), children with HLHS have an underdeveloped left ventricle that is unable to support the systemic circulation, along with hypoplasia of the ascending aorta and stenosis or atresia of the mitral and aortic valves (5). HLHS is no longer a fatal diagnosis because of the development of surgical palliation through 3-staged surgical operations (6–8). Nevertheless, HLHS has a high mortality of up to 36.5%, especially during the interstage period throughout the first year of life (9). This is due, at least in part, to the significant mechanical loads imposed upon the RV causing stress on the right ventricle (RV) that now must support systemic circulation. Increased load can lead to RV hypertrophy and dysfunction, and often requires heart transplantation due to RV failure. However, transplantation is limited by organ availability, limited organ lifespan, and less beneficial outcomes in patient with HLHS when compared with other indications (10).

The unique challenges posed by HLHS have necessitated the pursuit of novel therapies to improve outcomes for the single-ventricle population. Cell-based therapy has the potential to safely improve myocardial function and reverse maladaptive remodeling following injury (11–14). Moreover, such approaches have shown promise in treating various adult cardiac conditions (12, 13, 15–17), and along with preclinical animal studies modeling RV pressure overload (18), and acute ischemic injuries (19), provide strong rationale for the application to HLHS. There are few studies investigating the use of cell-based therapies in patients with HLHS (20–25), and none to date have studied the use of MSCs. Autologous cell-based therpaies carry the benefits of avoiding immune therapies; however, have a number of drawbacks, including logistic and economic constrainsts, feasibilitiy failures for manusfacturing, and often undiagnosed medical comordities and potential genetic disease contributors. In contrast, allogeneic MSCs circumvent these issues and have shown potential promise over autologous MSCs in the POSEIDON trial for dilated cardiomyopathy patients (26). Preclinical and clinical studies have demonstrated that allogenic MSCs are immunoprivileged, and thus tissue-type matching and immunosuppressive therapy are not needed (26). Since the MSCs can be administered by direct intramyocardial injection, this obviates the need for traversing the endothelial barrier such as would be required with intracoronary administration.

Here we present the results of the phase I ELPIS trial. The primary objective was to demonstrate the safety and feasibility of intramyocardial injection of MSCs during the Stage II bidirectional cavopulmonary anastomosis (BDCPA) surgery (27). To address limitations to noninvasively monitor transplanted cells in a repeatable, time-sensitive manner, we assayed circulating exosomes of transplanted MSCs and used computational modeling to evaluate the bioactivity and dynamics of the transplanted cells.

## METHODS

This study was under the oversight of an independent Institutional Review Board (Western IRB: Puyallup, WA) and Data Safety Monitoring Board (DSMB). The consent form, protocol, and all supporting documents were also reviewed by the IRBs of participating centers: University Maryland Medical Center, Cincinnati Children’s Hospital Medical Center, and Primary Children’s Medical Center. Methods were previously reported. (27), and briefly described as follows.

### Study Design and Patient Population

The patients in this report were enrolled under two studies: a formal phase I safety study using an allogeneic MSC formulation called Lomecel-B (Longeveron Inc.: Miami, FL. *N*=10; NCT03525418); and a second set of 4 patients enrolled in a run-in using allogeneic MSCs produced by the Interdisciplinary Stem Cell Institute (University of Miami Miller School of Medicine: Miami, FL. *N*=4; NCT02398604). The aims were to assess: 1) the safety and feasibility of intramyocardial allogeneic MSC injection in patients with HLHS during stage II palliation; 2) provisional, hypothesis generating phenotypic data to design future phase II studies, and 3) novel exosome/microRNA biomarker information to gain mechanistic insights. Patients fulfilling inclusion and exclusion criteria were offered enrollment in the studies, and similar inclusion and exclusion criteria were used for both protocols. Inclusion criteria required enrolled patients to have any type of HLHS requiring BDCPA surgery. Exclusion criteria included: having HLHS and a severely restrictive or intact atrial septum; patients undergoing the Norwood procedure who do not have HLHS; presence of significant coronary artery sinusoids; patients requiring mechanical circulatory support before surgery; evidence of arrhythmia requiring antiarrhythmic treatment prior to enrollment; unwillingness or inability of parent(s) or guardian(s) to comply with necessary follow-up(s); patients who are serum positive for HIV, hepatitis B surface antigen, or viremic hepatitis C; or who are unsuitable for inclusion in the study as determined by the investigator. Parents or guardians provided written consent before enrollment (Figure 1A). Enrolled patients then received baseline cardiac magnetic resonance imaging (CMR) before the stage II operation. At Stage II palliation, patients received intramyocardial injections of allogeneic MSCs and received follow-ups at 6- and 12-months after surgery (Figure 1A).

**Figure 1.**
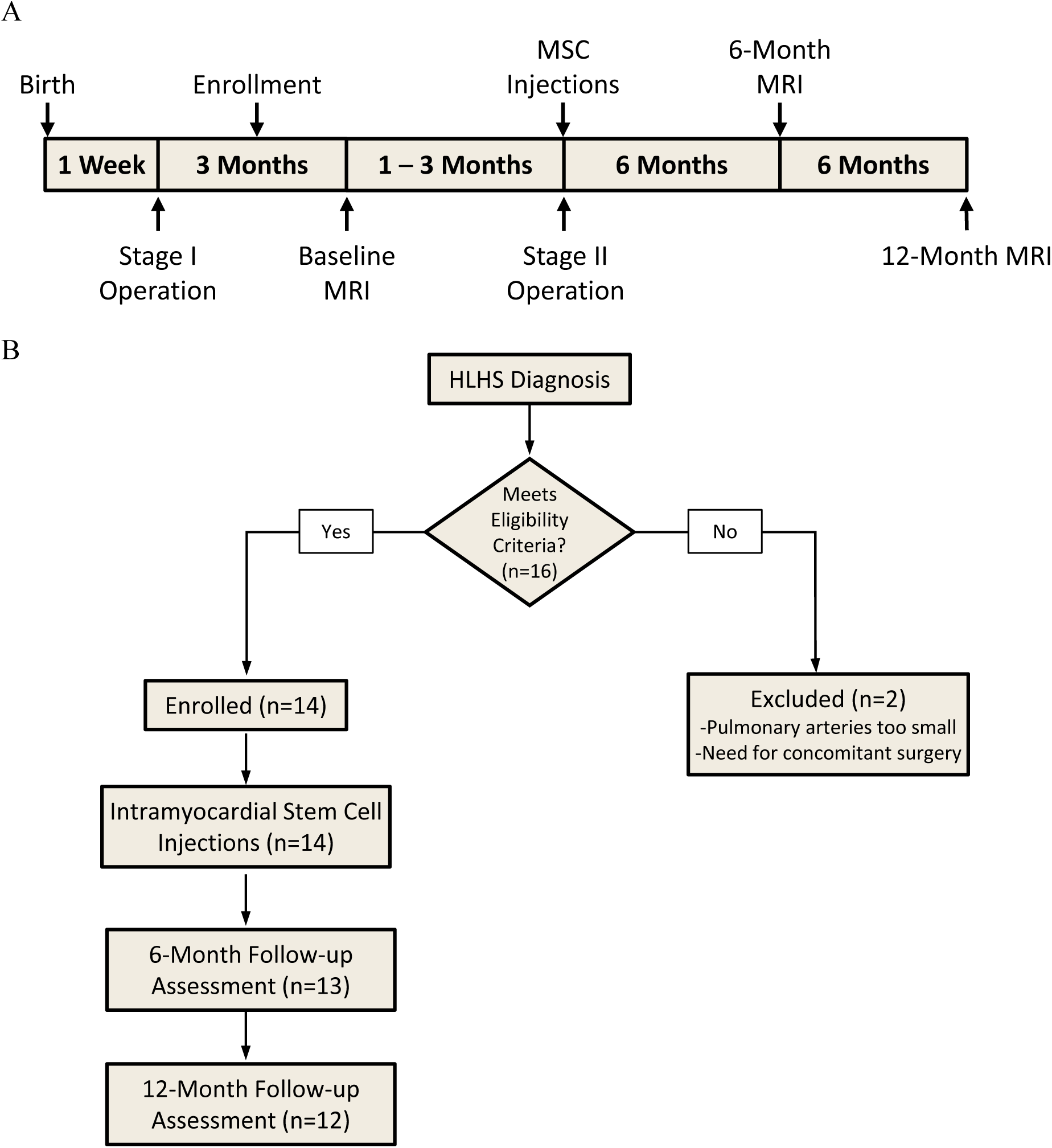
Study timeline and study flow, phase I of the ELPIS study. **A**, Study timeline. **B**, Study flow. Patients who met the eligibility criteria underwent BDCPA and intramyocardial injections, then were followed up with CMR at 6 and 12 months.

### Cell Harvesting and Intramyocardial Injection

Cells were produced using current Good Manufacturing Practices (12, 28) at Longeveron, Inc., Miami, FL (*N*=10; NCT03525418; Lomecel-B) and the University of Miami Interdisciplinary Stem Cell Institute (*N*=4; NCT02398604). The trials were conducted under two unique Investigational New Drug applications from the U.S. Food and Drug Administration. Cell harvesting, processing, and manufacturing were performed similar to as previously described (27). Release criteria for the MSCs met minimal release criteria of >70% viability, negative Gram staining, low levels of endotoxin, sterility, and being >95% CD105^+^ and CD90^+^ cells, consistent with previous trials (12, 28).

The allogeneic MSCs were delivered as previously described, and an RV map of the 8 injection sites in the RV free wall has been previously published (27). Target cell doses of 2.5 × 10^5^ cells/kg were used, actual doses based on weight in the formal phase I trial were between 1-2 x 10^6^ cells administered in 600 µL. The entire dose was divided and delivered in 4 intramyocardial injections of 100 µL and 4 intramyocardial injections of 50 µL per injection (27). The remainder of the BDCPA operation proceeded as anticipated. This was similar to the methods for intramyocardial administration and dosing of allogeneic MSCs used in the PROMETHEUS (28) and POSEIDON (12) trials.

### Study Endpoints

The primary objective of the phase I ELPIS trial was safety, with the primary endpoint being the incidence of MACE (defined as sustained/symptomatic ventricular tachycardia requiring inotropic support, aggravation of heart failure, myocardial infarction, unplanned cardiovascular operation for cardiac tamponade, or death) one-year post-BDCPA or infection within one month of treatment. The safety data was reviewed three times during the course of the trial and a final formal study report analysis was conducted by the independent DSMB. The DSMB did not report any concerns with safety or feasibility, and recommended advancement to the next phase.

Secondary endpoints included functional cardiac changes from baseline, change in somatic growth, and assessment of comorbidities. Cardiac function measurements included RV ejection fraction (RVEF), body surface area (BSA)-indexed RV stroke volume (RVSV), RV end-diastolic volume (RVEDV), RV end-systolic volume (RVESV), BSA-indexed RV end-diastolic diameter (RVDd), BSA-indexed RV end-systolic diameter (RVDs), RV global longitudinal strain (GLS), and qualitative tricuspid regurgitation (TR) and TR fraction (TR RF) measured by serial echocardiograms and CMR. Other secondary endpoints were need for transplantation, re-hospitalizations, cardiovascular mortality, and all-cause mortality.

### Exosome analyses

An exploratory sub-study was performed in 6 of the 14 patients in which exosome RNA expression was determined with Human Clariom S Assay and GeneChip miRNA 4.0 Arrays (Applied Biosystems), and correlated with functional outcomes. Exosomal miRNA was measured from 6 patients, and exosomal total RNA was profiled in 6 patients. Data were processed and annotated with the following R packages: oligo (29), pd.clariom.s.human (30), clariomshumantranscriptcluster.db (31), miRBaseVersions.db (32), and pd.mirna.4.0. (33) Lowly expressed RNAs and multiple mappings were removed. Then, data were quantile normalized with robust multi-array method. Based on plasma and CMR data availability, 6 months-to-1 year fold change values for RV mass, RVEDV, RVESV, RVSV, RVEF, TR RF, and GLS from 6 patients (E1, E2, E3, E4, E5, E6) were used in subsequent analyses.

### Unsupervised analysis of filtered, normalized RNA

Differential expression of gene (DEG) analysis for day two and day seven patient matched exosomes was performed using a paired, two-tailed Student’s t-test. Results indicated similar expression levels at both days (0.10% of RNAs with p<0.05 and |fold change| >1.5). Thus, for the remaining analyses, day seven exosome RNA expression was used. Principal component analysis (PCA) was performed using the SIMCA-P software (Umetrics, Sartorius Stedim Biotech).

The WGCNA R package was used to construct co-expression networks for the filtered, normalized genes. The details of this algorithm are described by Langfelder and Horvath(31). Briefly, the optimal soft-threshold power was graphically determined (β = 12 and 20 for miRNA and RNA) and the minimum module size was set to 50. Clusters, or modules, of RNAs were determined by first computing the adjacency matrix and then transforming it to form the topological overlap matrix (TOM). Then, the corresponding dissimilarity matrix, 1-TOM, and the cutree Dynamic function was used for hierarchical clustering and module detection. Highly correlated modules (r > 0.75) were merged to form the final co-expression modules. Twenty-five miRNA and RNA modules were determined, each. The dissimilarity of the module Eigengenes was computed with the module Eigengenes function and the association between Eigengene values and CMR outcomes were assessed by Spearman’s correlation.

### Partial least squares regression (PLSR) and biological pathway analyses

PLSR modeling was performed using SIMCA-P software (Umetrics, Sartorius Stedim Biotech). A regression model was built from six patients’ (E1, E2, E3, E4, E5, E6) day seven exosome RNA signals, with 6 months-to-1 year fold change CMR values. Feature selection was performed to select the top 200 RNA signals with the highest variable importance in the projection (VIP) values and a final two-component model was built. miRTarBase was used to identify experimentally validated miRNA gene targets (≥ 3 experiments; mirtarbase.cuhk.edu.cn). Gene Ontology and KEGG pathway enrichment of genes and miRNA gene targets was determined using Metascape (metascape.org). The web-based Metascape tool determined the significantly enriched terms from gene sets (p<0.05).

### Statistical Analyses

Cardiac functional parameters, somatic growth, and brain natriuretic peptide (BNP) were compared at the various timepoints. Normality of parameter distributions was evaluated using a Kolmogorov-Smirnov test. For continuous measurements that were not normally distributed at all timepoints, comparisons among values at the 3 timepoints were made using a Kruskal-Wallis H-test followed by a Dunn’s Multiple Comparison test. Normally distributed measurements were compared using one-way analysis of variance (ANOVA), with correction for multiple comparisons. The statistical significance threshold was *p* ≤ 0.05. Statistical analyses were completed using SPSS and GraphPad Prism 9.

## RESULTS

### Patient and Operative Characteristics

This report comprises the formal phase I ELPIS trial (*N*=10), and an additional group (*N*=4) with similar characteristics enrolled as a run-in. The baseline characteristics of the patients are shown in Table 1. All participants were diagnosed with HLHS, with most patients having mitral and aortic atresia (61.5%). The average weight for age z-score was -0.9 for the study group. Each patient had previously undergone the Stage I (Norwood) surgery with a Sano shunt at an average age of 4.4 days (Table 2).

**Table 1.** Baseline Demographics

| Characteristic<sup>a</sup> | Lomecel-B<br>(N=10) | Allo-MSCs<br>(N=4) | All (N=14) |
| --- | --- | --- | --- |
| Age at BDCPA operation (months) | 4.89±0.85 | 4.83±0.59 | 4.87±0.76 |
| Male gender | 7 (70.0) | 1 (25.0) | 8 (57.19) |
| Ethnicity |  |  |  |
| Hispanic or Latino | 0 | 2 (50.0) | 2 (14.3) |
| Not Hispanic or Latino | 10 (100.0) | 2 (50.0) | 12 (85.7) |
| Race |  |  |  |
| White | 7 (70.0) | 0 | 7 (50.02) |
| Black/African American | 3 (30.0) | 2 (50.0) | 5 (35.7) |
| Not collected | 0 | 1 (25.0) | 1 (7.1) |
| Other | 0 | 1 (25.0) | 1 (7.1) |
| Body Surface Area (m<sup>2</sup>) | 0.331±0.03 | 0.327±0.032 | 0.33±0.029 |
| Length for Age Z-Score | -0.87±2.02 | -1.350±0.719 | -1.01±1.73 |
| Weight for Age Z-Score | -1.09±1.31 | -0.450±0.656 | -0.91±1.17 |
| Duration of Bypass (min) | 113.1±17.44 | 101.0±11.14 | 109.6±16.69 |
| Duration of Injection (min) | 5.5±1.78 | 6.2±0.96 | 5.7±1.59 |
| Hospital Length of Stay (days) | 11.7±9.58 | 7.25±0.50 | 10.43±8.24 |
| Norwood Shunt Type |  |  |  |
| RV-PA (Sano) | 10 (100) | 4 (100) | 14 (100) |
| RV Function by CMR |  |  |  |
| RV Ejection Fraction (%) | 62.62±5.99 | 67.97±11.02 | 64.15±7.69 |
| RV End Systolic Volume Index (mL/m<sup>2</sup> BSA<sup>1,3</sup>) | 49.7 1± 15.61 | 61.38±48.43 | 53.04±27.2 |
| RV End Diastolic Volume Index (mL/m<sup>2</sup> BSA<sup>1,3</sup>) | 133.59±36.14 | 170.25±85.19 | 144.06±53.62 |
| RV Stroke Volume Index (mL/m<sup>2</sup> BSA<sup>1,3</sup>) | 83.88±23.65 | 108.87±37.03 | 91.02±29 |
| RV Mass Index (g/m<sup>2</sup> BSA<sup>1,3</sup>) | 101.26±33.71 | 116.15±34.97 | 105.52±33.43 |
| Tricuspid regurgitant fraction | 0.45±0.19 | 0.51±0.20 | 0.48±0.18 |
| RV systolic diameter (mm/m<sup>2</sup> √BSA) | 37.5±9.24 | 40.49±22.07 | 38.35±13.17 |
| RV diastolic diameter (mm/m<sup>2</sup> √BSA) | 61.36±13.95 | 68.00±21.56 | 63.26±15.87 |
| GLS (%) | -24.39±6.99 | -21.29±6.65 | -23.26±6.71 |
| Sphericity Index | 1.31±0.35 | 1.30±0.26 | 1.30±0.31 |

<sup>a</sup>Values are n (%) or mean±SD unless otherwise noted

**Table 2.** Patient characteristics at the time of stage II procedure

| Characteristic<sup>a</sup> | Lomecel-B<br>(N=10) |
| --- | --- |
| Anatomic diagnosis HLHS | 10 (100.0) |
| Morphology |  |
| Mitral atresia and aortic atresia | 6 (60) |
| Mitral stenosis and aortic atresia | 3 (30) |
| Mitral atresia and aortic stenosis | 0 |
| Mitral stenosis and aortic stenosis | 1 (10) |
| Partially restrictive atrial septal defect | 1 (10) |
| Sinusoidal arteries | 0 |
| Aortic size before first palliation (mm) | 1.70±0.46 |
| Cardiac position levocardia | 10 (100.0) |
| Associated anomaly |  |
| Right aortic arch | 0 |
| Bilateral SVC | 1 (10) |
| Previous cardiac surgical procedures |  |
| Norwood operation | 10 (100.0) |
| Age at Norwood (days) | 4.3±1.25 |
| Modified Blalock-Taussig shunt | 0 |
| Sano shunt | 10 (100.0) |
| Cardiac arrest | 0 |
| ECMO | 0 |
| History of bilateral pulmonary artery banding | 1 (20) |
| TAPVC | 1 (10) |
| Damus-Kaye-Stansel anastomosis | 3 (30) |
| History of catheter interventions |  |
| Balloon atrial septostomy | 1 (10) |
| APCA coil occlusion | 1 (10) |
| Pulmonary artery angioplasty | 0 |
| Balloon of distal coarctation of aorta | 2 (20) |
| Preoperative assessment |  |
| Genetic syndrome | 0 (0) |
| Extracardiac malformation | 1 (10) |
| Mild tricuspid regurgitation | 1 (10) |
| Oxygen saturation % | 80.6±3.34 |
| Aortic saturation % (n=9) | 78.06±10.01 |
| Mean BNP [range] (µg/mL) | 759.1 [45.0 – 3190.0] |
| Mean pulmonary artery pressure (mmHg) (n=9) | 13.56±2.55 |
| Cardiac index (L/min/m<sup>2</sup>) (n=9) | 3.25±0.8 |
| Coarctation of aorta (n = 7) | 1 (10) |
| Medication profile |  |
| Diuretics | 7 (70) |
| Digoxin | 10 (100) |
| Digitalis | 0 (0) |
| ACE inhibitor or ARB | 4 (40) |
| PDE5 | 0 |
| Endothelin-1 receptor antagonist | 0 |
| Aspirin (Antiplatelet) | 10 (100) |
| Beta-blocker | 2 (20) |
| Antiarrhythmic agent | 0 (0) |
| Anti-reflux agent | 8 (80) |

<sup>a</sup>Values are n (%) or mean±SD unless otherwise noted
ACE, Angiotensin-converting enzyme; APCA, aortopulmonary collateral arteries; ARB, angiotensin II receptor blocker; BNP, brain natriuretic peptide; ECMO, extracorporeal membrane oxygenation; HLHS, hypoplastic left heart syndrome; PDE5, phosphodiesterase type 5; SVC, superior vena cava; TAPVC, total anomalous pulmonary venous connection

All patients received intramyocardial MSC injections during the BDCPA procedure. The total cardiopulmonary bypass averaged 113.1±17.44 minutes (Table 3). The average time for allogeneic MSC injection was 5.5±1.78 minutes. No patient underwent aortic cross-clamping or deep hypothermic circulatory arrest. Concomitant atrial septectomy was completed in three patients and concomitant pulmonary artery patch augmentation was completed in four patients; the remaining patients underwent no concomitant procedures. Nine of the patients enrolled in the formal phase I trial were seen at the 6-month follow-up and 8 were seen at the 12-month follow-up (Figure 1B).

**Table 3.** Operative characteristics of study population during bidirectional cavopulmonary anastomosis and stem cell injection

| Operative Variable<sup>a</sup> | Lomecel-B(N=10) |
| --- | --- |
| Concomitant procedures |  |
| Atrial septectomy<sup>a</sup> | 4 (40) |
| Pulmonary artery patch augmentation<sup>a</sup> | 4 (40) |
| Aortic cross clamp |  |
| Number of patients | 0 |
| Average time (min) | 0 |
| Deep hypothermic circulatory arrest |  |
| Number of patients | 0 |
| Average time (min) | 0 |
| Average MSC trial injection time (min) | 5.5±1.78 |
| Average volume of MSC injected (µL) | 600.0±0.0 |
| Average number of MSC injections | 8.0±0.0 |
| Average length of hospitalization (days) | 11.7±9.58<sup>b</sup> |

<sup>a</sup>Values are N (%) or mean±SD unless otherwise noted
<sup>b</sup>Length of stay consistent with 2016-2020 STS Benchmark data (unpublished) which shows median postoperative length of stay for Glenn/hemi-Fontan procedures is 17.8 days.
MSC, medicinal signaling cell

### Safety: Results of the formal phase I study

Ten patients successfully underwent BDCPA receiving intramyocardial Lomecel-B injection during the Stage II surgery. No adverse events on trial were attributed to Lomecel-B (Table 4). Specifically, no treated patients had myocardial infarction, death, ventricular arrythmia, systemic infection, stroke, or allergic reaction. No myocardial ischemia was seen by monitoring electrocardiogram and telemetry. Two patients experienced an infectious event within one month of surgery: one urinary tract infection; and one methicillin-resistant Staphylococcus aureus bacteremia. These events were not considered related to treatment. No mortality or heart-transplantion was recorded in the one year following treatment. Ten re-hospitalizations occurred in 5 patients, which were not related to worsening RV function. One subject required catheter-mediated angioplasty for an ascending aortic obstruction. HLA class I reactions were decreased at 26 weeks post-treatment as compared to baseline (*p* = 0.031; Figure 4E). HLA class II reactions showed a trend toward reduction (*p* = 0.25, Figure 4F).

**Table 4.** Incidence of major adverse cardiac events and co-morbidities.

|  | Number of Incidences |
| --- | --- |
|  | Lomecel-B (N=10) |
| Major Adverse Cardiac Events<sup>a</sup> |  |
| Sustained/symptomatic ventricular tachycardia requiring intervention with inotropic support | 0 |
| Aggravation of heart failure | 0 |
| Myocardial infarction | 0 |
| Unplanned cardiovascular operation for cardiac tamponade | 0 |
| Death | 0 |
| Co-morbidities |  |
| Cardiovascular morbidity<sup>b</sup> | 1 |
| Need for transplantation | 0 |
| Re-hospitalizations | 10 (5 patients) |
| Cardiovascular mortality | 0 |
| All-cause morbidity | 0 |

<sup>a</sup>Evaluated through 1 year post-treatment
<sup>b</sup>The one cardiovascular morbidity was an ascending aorta obstruction which required angioplasty

### Postoperative Outcomes

Cardiac function was assessed by CMR 3 times over a 1-year period after treatment. RVEF which was normal at baseline (62.62±5.99%) showed a non-significant trend towards reduction at 6 and 12 months (53.69±9.56% and 52.31±5.63%, respectively) (Figure 2A, Supplemental Tables 1 and 2). RVSV index did not significantly change from baseline to 6 months (−4.05±33.35 ml, *p* > 0.05) or 12 months (−12.75±34.8 ml, *p* > 0.05; Figure 2B, Supplemental Table 1). RVDd and RVDs did not significantly change from baseline at either 6 or 12 months (*p* > 0.05, Figure 2C, 2D, Supplemental Table 1). GLS did not change from baseline to 6- or 12-months post-treatment (Figure 2E, Supplemental Table 1). TR was assessed subjectively and quantitatively by CMR (Figure 3A, 3B). The mean TR RF decreased from 0.45±0.19 at baseline to 0.32±0.06 (*p* > 0.05), and to 0.06±0.09 (*p* > 0.05; Figure 3B, Supplemental Table 1). The serum BNP increased following day two post-treatment, decreased following week twenty-four post-treatment (Figure 4A) and the change in BNP from baseline was not significant (*p* > 0.05; Figure 4B). No significant changes were observed in weight for age z-score at 6 months (*p* = 0.30 vs baseline) or 12 months (*p* = 0.73 vs baseline; Figure 4C, Supplemental Table 1). No significant changes were observed in length for age z-score at 6 months(*p* = 0.079 vs baseline) or 12 months (*p* = 0.946 vs baseline; Figure 4D, Supplemental Table 1). However, the length for age z-score significantly decreased from 1.24±1.64 at 6-month to -0.40±1.83 at 12-months (*p* = 0.0136).

**Figure 2.**
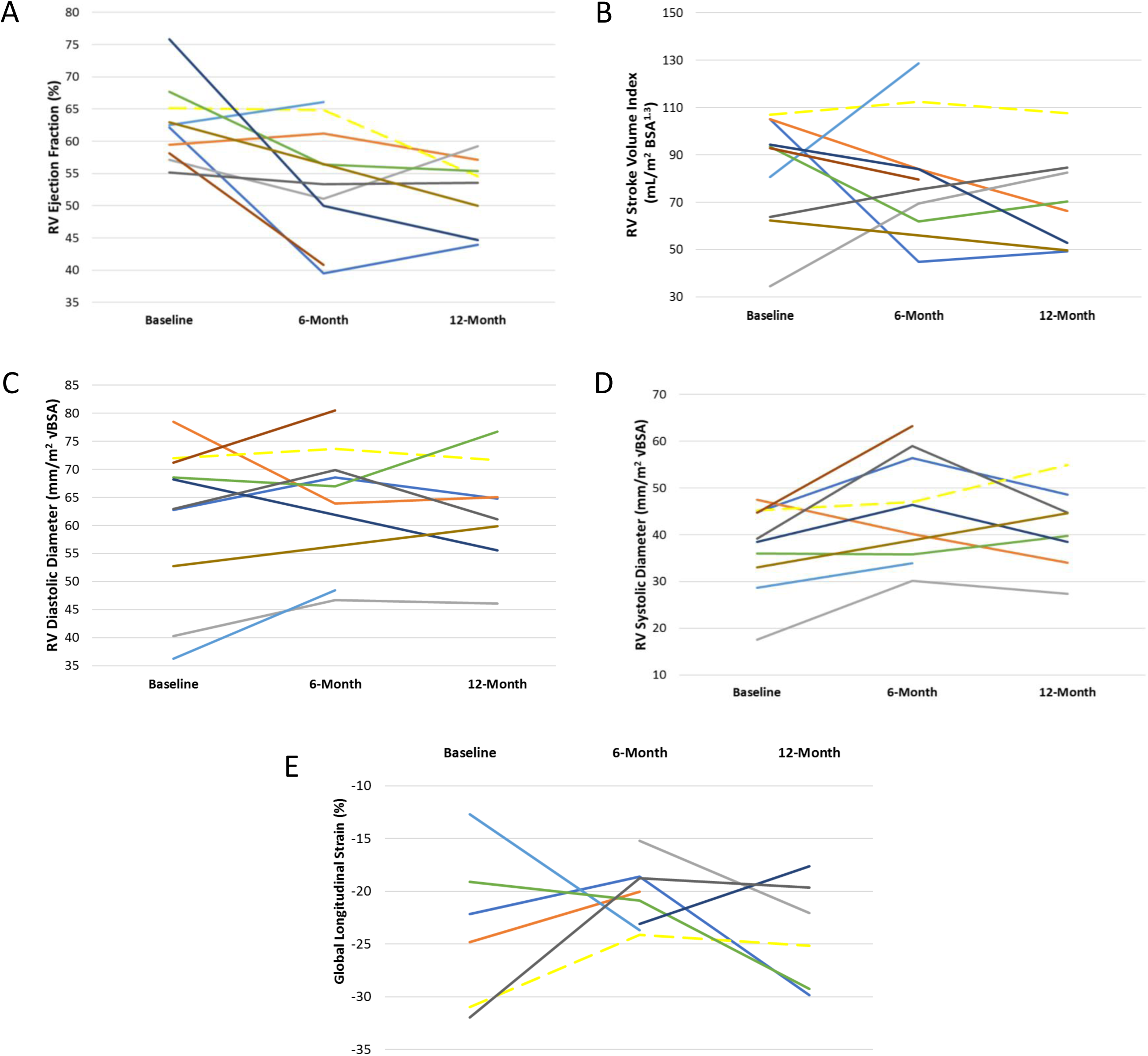
Cardiac function at baseline, 6 months follow-up, and 12-month follow-up in Lomecel-B treated patients (*N*=10). **A**, Right ventricular ejection fraction did not change over the follow-up period compared to baseline. **B**, Right ventricle stroke volume index did not change over the follow-up period compared to baseline. **C**, Right ventricle diastolic diameter did not change over the follow-up period compared to baseline. **D**, Right ventricle systolic diameter did not change over the follow-up period compared to baseline. **E**, Global longitudinal strain did not change over the follow-up period compared to baseline.

**Figure 3.**
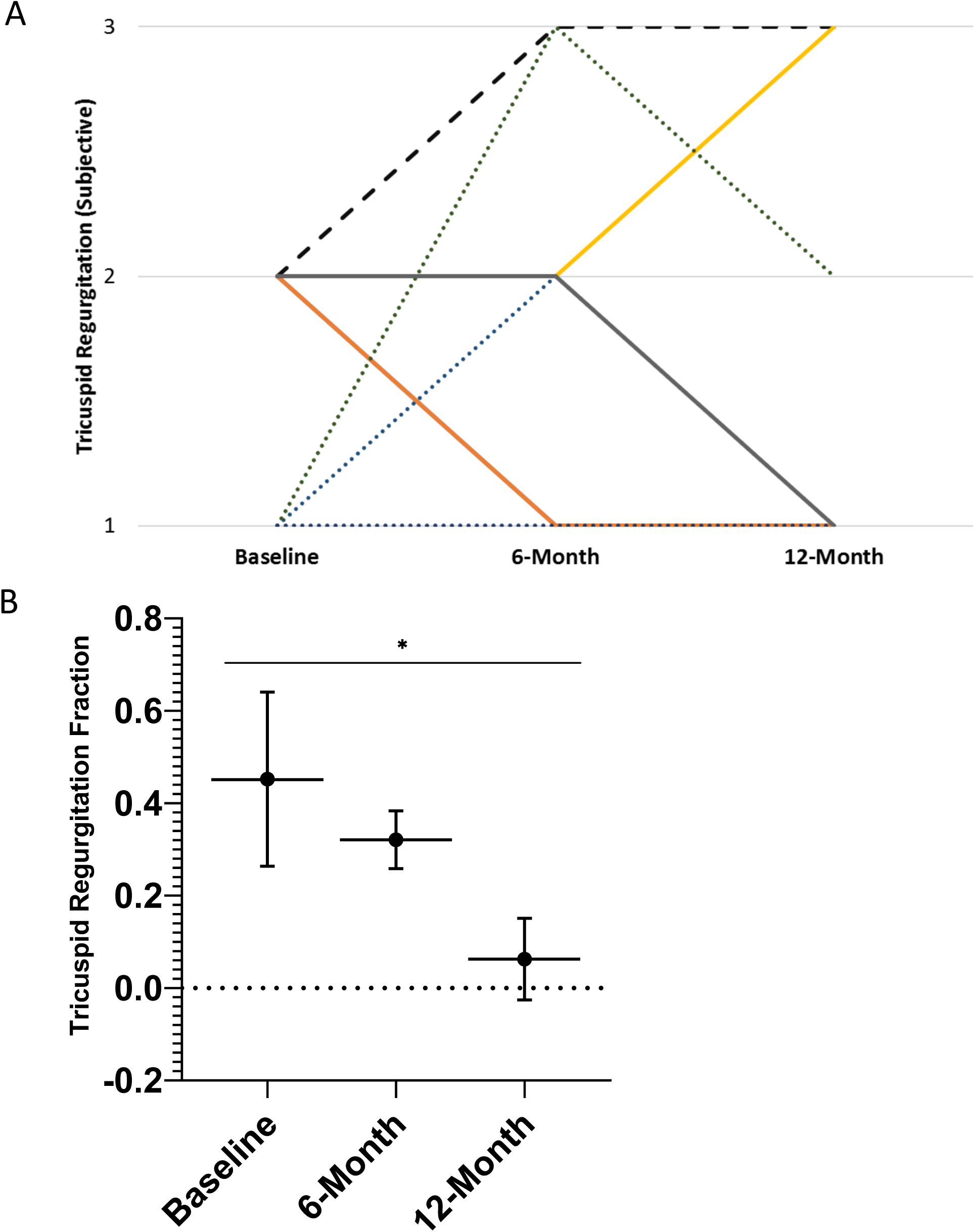
Subjective tricuspid regurgitation at baseline, 6-month, and 12-month follow-up. **A**, Subjective regurgitation was quantified as severe (3), moderate (2), or mild (1). **B**, Tricuspid regurgitant fraction was measured. Reported as mean +/- standard deviation. * indicates p < 0.05.

**Figure 4.**
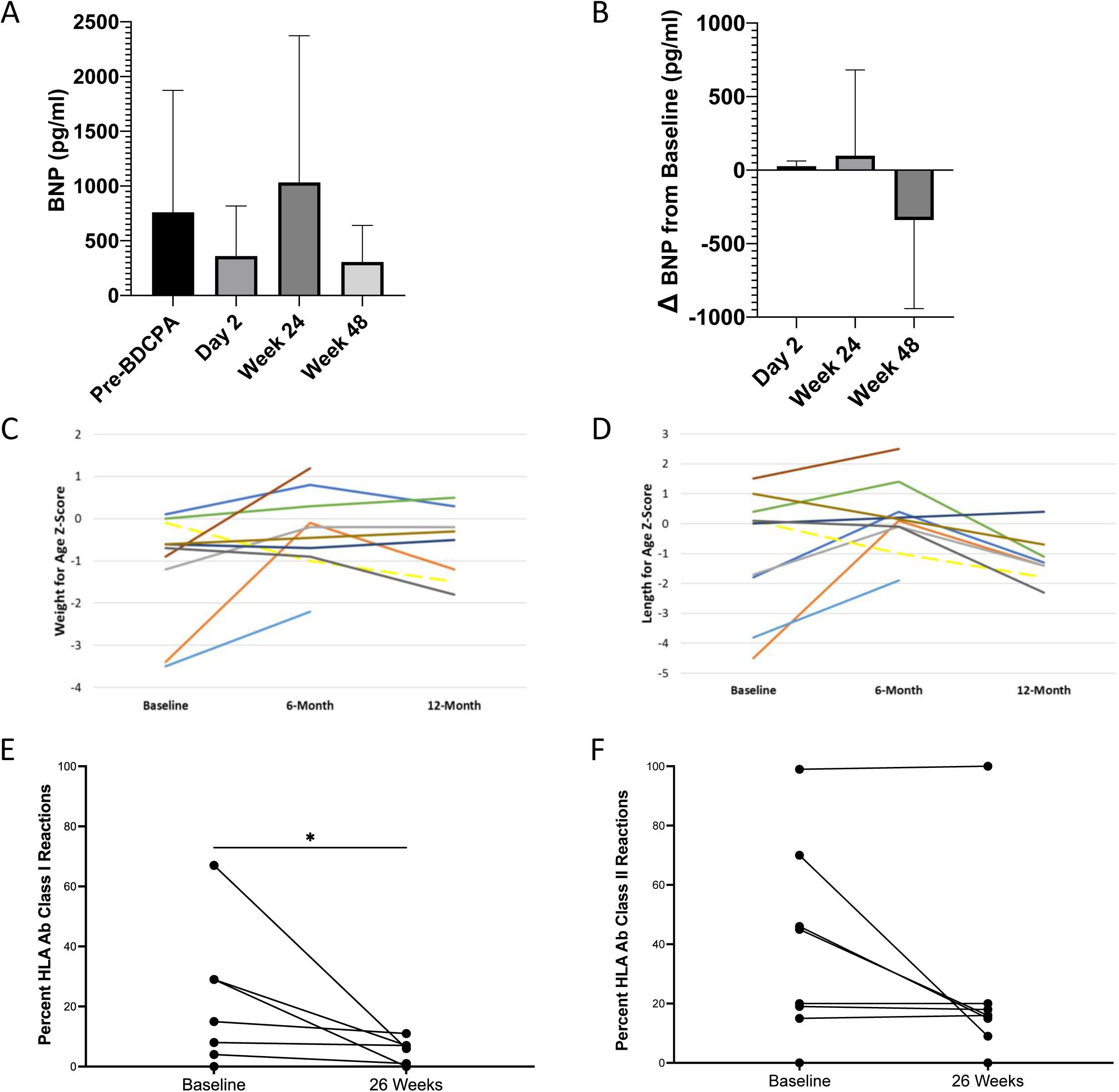
Baseline and follow up data monitoring patients throughout ELPIS phase I trial Lomecel-B treated patients. **A-B**, Serum BNP increased following day two post-treatment but decreased following week twenty-four post-treatment. No changes were statistically significant. Error bars indicate standard deviation. **C**, Somatic growth of patients at baseline and at follow-up. Weight for age z-score increased from baseline to 6 months. Weight for age z-score did not differ between baseline and 6 or 12 months. **D**, Length for age z-score increased from baseline to 6 months. The length for age z-score significantly decreased from 6-month to 12-months (p = 0.0136) **E**, HLA class I reactions significantly decreased from baseline to 26 weeks post-treatment. **F**, HLA class II reactions showed a trend towards decreasing after treatment, but this did not reach statistical significance. Only patients with HLA data at baseline and 26 weeks are represented (*N*=8). * indicates *p* < 0.05. BNP, brain natriuretic peptide. .

### RNA expression profiles in MSC-derived exosomes

We hypothesized that injected MSCs would release exosomes composed of allogeneiccell-specific constituents, and that these would be detectable in the recipient blood. Using a MHC mismatch technique based on the donor and recipient MHC profiles, MSC-derived exosomes (HLA-I+) were collected at 2 and 7 days post-treatment from the plasma of 6 patients: 2 from the phase I safety study, and 4 from the run-in (E1-E6). Immunoblot analysis demonstrated allogeneic MSC-derived exosomes expressed respective HLA-I molecules, flotillin (exosome marker), and c-kit (progenitor marker) (Figure 5A, Figure 6). Total RNA and miRNA in exosomes were quantified by microarray, and differential expression of RNAs between time points were assessed (Figure 5B). Exosomal expression profiles remained stable between days 2 and 7 with 0.1% (17 of 16,351) of RNAs differing (p < 0.05 and |fold change| > 1.5). Additionally, principal component analysis (PCA) of RNA and miRNA arrays do not show distinct clustering of day 2 and 7 samples (Figure 5C, 5D). However, there was separation of patients E2 and E3 in total RNA PCA plot.

**Figure 5.**
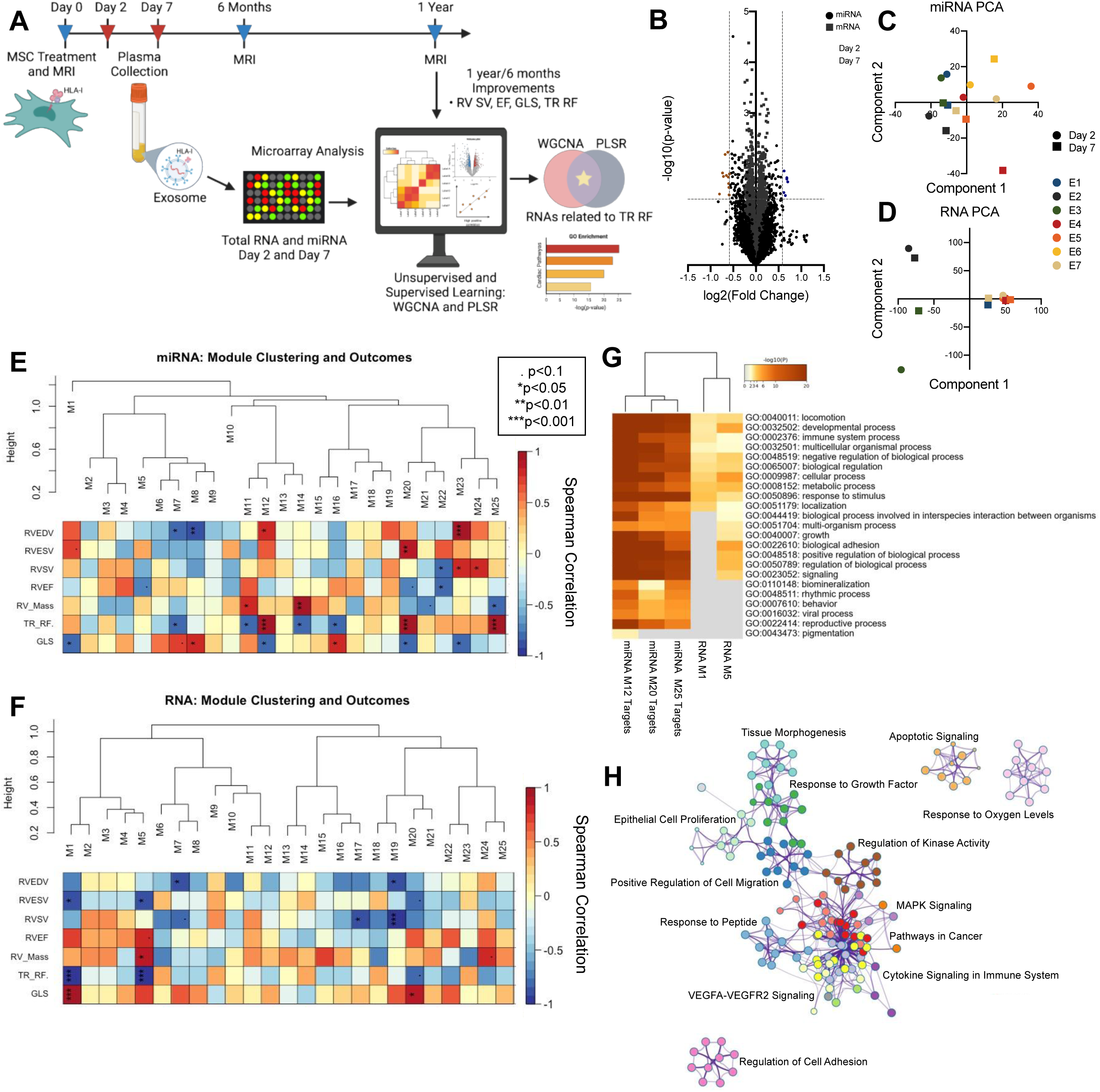
Unsupervised analysis of MSC-derived exosome RNA. **A**, Plasma exosomes from days 2 and 7 were assessed for miRNA and total RNA content with microarray. RNA expression was related to CMR 6 months-to-1 year fold change values. **B**, Volcano plot of RNA expression fold change from days 2 to 7. Few miRNAs (circles) and RNAs (squares) upregulated in days 2 and 7 are highlighted in orange and blue, respectively. Principal component analysis of days 2 and 7 miRNAs (**C**), and RNAs (**D**). Merged dynamic clustering from weighted correlation network analysis (WGCNA) detected 25 modules of highly correlated day 7 miRNAs (**E**) and RNAs (**F**). Module correlation to CMR outcomes (6 months-to-1 year fold change) are depicted in heatmaps (red = high, positive correlation). Modules significantly correlated to an CMR outcome (p<0.1) are marked. **G,** Enriched gene ontology (GO) parent terms of WGCNA RNA modules negatively correlated to TR RF (M1 and M5) and gene targets of miRNA modules positively correlated to TR RF (M12, M20, M25). **H,** A network of experimentally validated gene targets of miRNAs positively correlated to TR RF (M12, M20, M25). Enriched terms are colored by cluster (Metascape).

**Figure 6.**
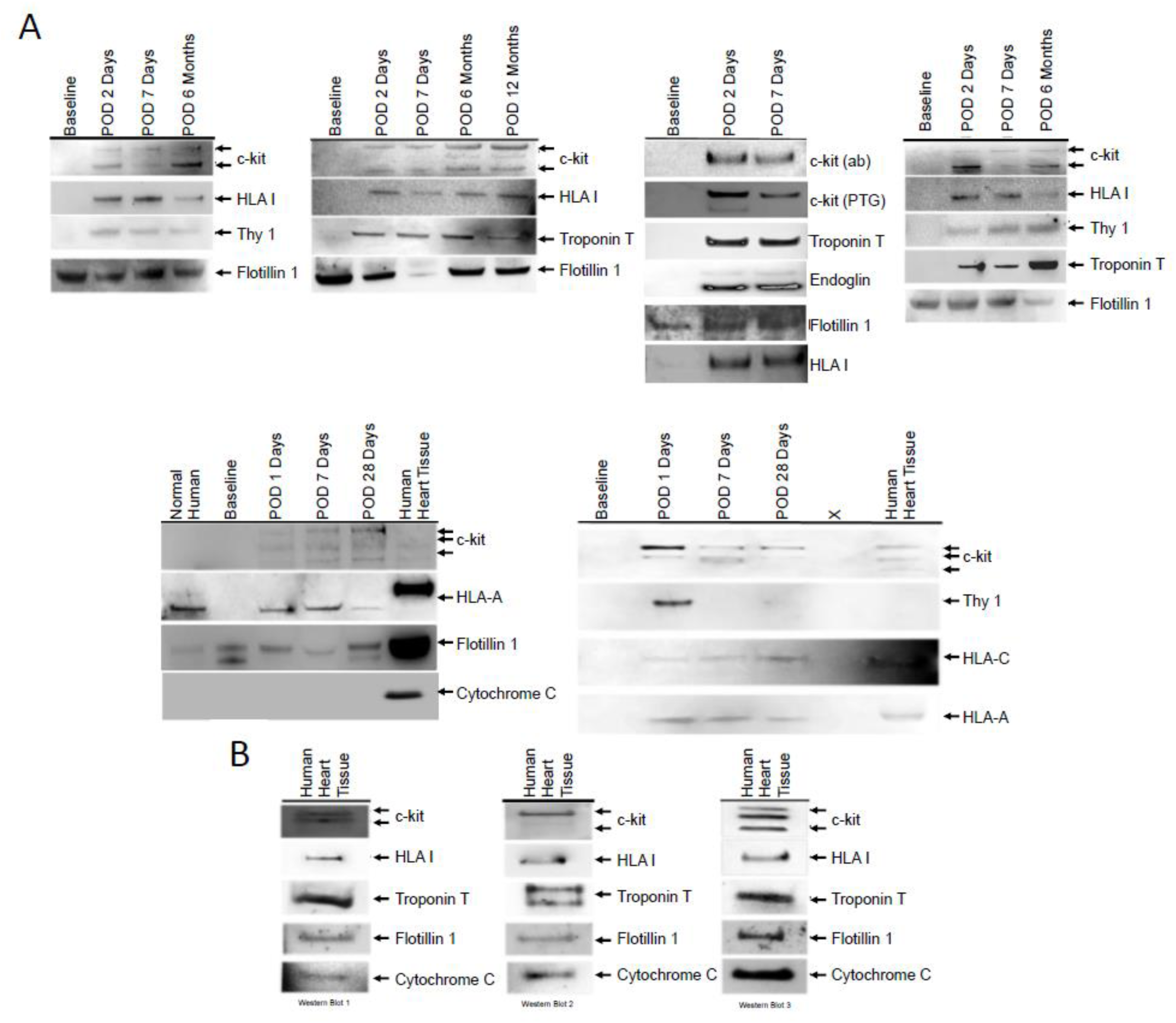
Representative western blots **A,** donor stem cell exosomes and **B**, tissue positive controls. POD, post-operative day.

CMR endpoints (RVSV, RVEDV, RVESV, RV mass, RVEF, GLS, and TR RF) were collected six-months and one-year after surgery and MSC treatment from six patients: E1, E2, E3, E4, E5, E6. TR RF was not collected for E5 and E6. Based on CMR and plasma availability, further analyses were conducted with these patients (*N*=6), using fold change values, or improvements, from six-months to one year.

### Weighted correlation network analysis (WGCNA) of day 7 exosomes

WGCNAwas performed with day 7 exosomal total RNA and miRNA expression values to construct co-expressed networks and identify co-expression clusters, or modules. Twenty-five co-expression modules were identified for both total RNA and miRNA, and their hierarchical clustering is illustrated in the dendrograms (Figure 5E, 5F). Modules of highly correlated genes were determined independently from CMR outcomes. Then, modules were summarized by module eigengenes and related to CMR 6 months-to-1 year fold change values in the lower heatmaps. Modules that significantly correlated with an outcome, negatively or positively, are marked (*p* < 0.1).

Next, we investigated the exosomal RNA modules which correlated with TR RF improvement, or six-months to one-year reductions. Three miRNA modules (M12, M20, and M25) positively correlated with TR RF fold change (r < 0, *p* < 0.001), and two total RNA modules (M1 and M5) negatively correlated with TR RF fold change (r > 0, *p* < 0.001). To determine the biological significance of these modules, we first identified experimentally validated miRNA gene targets, and then completed pathway analysis on the miRNA module gene targets and total RNA module genes. Enriched gene ontology parent terms of the modules include immune system processes, cellular processes, biological adhesion, and growth (Figure 5G). Pathway analysis of the three miRNA modules’ gene targets alone indicate enrichment of terms including, positive regulation of cell migration, response to growth factor, VEGFA-VEGFR2 signaling, and apoptotic signaling (Figure 5H).

### Partial least squares regression (PLSR) of day 7 exosomes

In addition to using unsupervised WGCNA, we also conducted supervised analysis with PLSR to establish a relationship between exosomal RNAs and CMR outcomes. A regression model was built from six patients’ (E1, E2, E3, E4, E5, E6) day seven exosome RNA signals, with 6 months-to-1 year fold change CMR values. Feature selection reduced the initial list of 16,351 to the 200 RNA variables with the greatest importance for the model projection (VIPs). Notably, the resulting feature reduction increased the proportion of miRNAs considered, indicating their importance in the regression model (Figure 7A). A two-component model of these top 200 VIPs captured the variance of RNA signals and CMR outcomes with high coefficients of determination: R^2^(RNA) = 0.927, R^2^(CMR Outcomes) = 0.865. The PLSR scores plot shows the spread of patient samples across the two components (Figure 7B). The PLSR loadings plot displays how the 200 VIP signals covary with CMR outcomes and show two distinct clusters across component one (Figure 7C). Furthermore, the top 200 RNA signals predicted CMR outcomes with high R^2^ values, slopes ∼1, and low root mean square error of estimation (Figure 7D).

**Figure 7.**
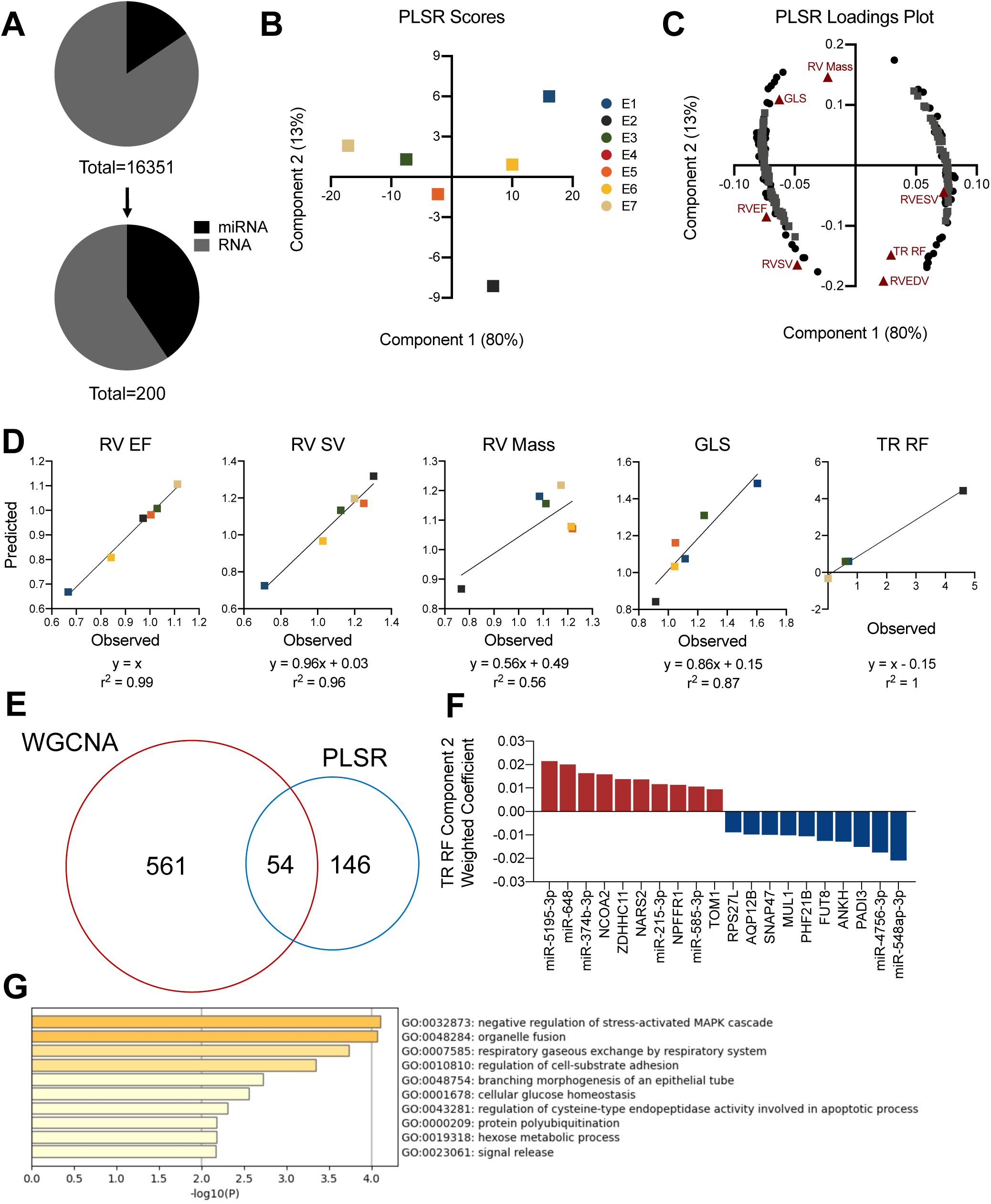
Partial least squares regression (PLSR) modeling of day 7 exosome RNA and identification of RNA signals related to TR RF. A two-component model was trained using the top 200 RNA variables of importance for the model projection (VIPs): R2(RNA) = 0.927, R2(CMR Outcomes) = 0.865. **A**, Total number of RNAs in the model was reduced to 200 from the initial set of 16,351. A higher proportion of the VIP RNAs are miRNAs. **B**, Scores plot of components 1 and 2 from the PLSR model trained with day 7 top 200 VIP RNAs. Components 1 and 2 explain 80% and 13% of the the variance, respectively. **C**, Loadings plots of components 1 and 2 from the PLSR model show the VIP RNAs covarying with CMR outcomes. **D,** PLSR model predictions of CMR outcomes correlated with overserved measurements. RNAs related to TR RF were determined from WGCNA and PLSR analyses. **E,** Venn diagram of WGCNA modules significantly correlated to TR RF (p<0.001, miRNA M12, M20, M25 and RNA M1, M5) and PLSR 200 VIP RNAs. 54 TR RF-related RNAs identified from the intersection. **F,** Top 10 positive and negative weighted coefficients from TR RF-related RNAs were determined with PLSR. **G,** Pathway analysis of 54 TR RF-related RNAs (Metascape).

### Intersection of Unsupervised and Supervised Learning Methods - RNAs related to TR RF

To determine the RNAs related to changes in TR RF, we considered the intersection of both analyses, WGCNA and PLSR. We overlapped genes from the WGCNA modules which correlate to TR RF fold change (miRNA modules M12, M20, M25 and total RNA modules M1 and M5) and the top 200 VIPs from the PLSR module (Figure 7E). We identified 54 RNAs in the intersection and determined the RNAs with the largest weighted coefficient for TR RF fold change in the PLSR model (Figure 7F). Pathway analysis of these 54 RNAs indicated significant enrichment of terms including, negative regulation of stress-activated mitogen-activated protein kinase cascade, regulation of cell-substrate adhesion, and apoptotic processes (Figure 7G).

## DISCUSSION

Our results suggest that MSCs can be safely and feasibly injected into the myocardium during cardiac surgery for HLHS patients. Injection of allogeneic MSCs did not lead to apparent adverse effects for up to 1 year, thus fulfilling the trial objectives. In addition, detailed CMR phenotyping was undertaken to provide hypothesis generating data for larger trials aimed at further delineating the effects of Lomecel-B on systemic RV performance. These studies provide key findings suggesting that Lomecel-B may improve tricuspid valve function and RV global longitudinal strain, possible evidence of improved ability to withstand the increased loads imposed by placing the RV in the systemic position. Additionally, RNAs were identified in the serum specific exosomes which provide a liquid biopsy that may be clinically useful in the development of this approach, and may offer mechanistic insights into potential MSC effects that could contribute to positive remodeling of the myocardium in HLHS patients.

MSCs are particularly attractive for pediatric applications because they can be reproducibly isolated and expanded from donor bone marrow (34, 35). These cells have immunosuppressive and pro-vascular properties and are effective as an allogeneic cell type as seen in previous adult clinical trials (36–40), making them strong therapeutic candidates.

This trial enrolled patients with normal RVEF at baseline. Consistent with previous natural history studies, RVEF drops following the stage II BDCPA due to loading condition alterations, but remains in the normal range (41). It is estimated that∼10% of children develop frank RV failure and require transplantation or die, with less than half of the children having transplant free survival to 15 years of age (42). Accordingly, the initial ELPIS results are consistent with the salutary effects of MSCs reported in preclinical studies, and are similar to the results of a variety of non-cardiac stem cells or progenitor cells used previously in single-ventricle patients (20–22, 25). At the 3-year follow-up of a phase II clinical trial, autologous intracoronary injection of cardiosphere-derived cells was a safe therapy and improved RVEF in single-ventricle patients with severe RV dysfunction at either their second or third palliative surgeries; however, the small, mixed study population of different staged single-ventricle patients limited the ability to draw definitive conclusions (20). Another phase I clinical trial demonstrated the safety of intramyocardial injections of autologous umbilical cord-derived mononuclear cells in HLHS patients, but no improvement of favorable RV remodeling was reported (24). Our results take advantage of an allogeneic cell type with the additional benefits of using a potentially “off-the-shelf”, non-time constrained product which can be frozen and used with repeat dosing for optimal efficacy.

We elected to deliver MSCs at the second staged operation to allow for stabilization of the single-ventricle physiology in HLHS patients at a time when the mortality rate is lower compared to the period between the first and second operations. There was no mortality in this small population of treated HLHS patients, with a 100% one-year transplant-free survival rate. In addition, our results demonstrate a non-statistical trend toward a reduction in RVEF from baseline to 6- and 12-months post-treatment. Indeed, the observed trend is consistent with previous published data reporting a decrease in RVEF from 58.3±9.1% at stage I of HLHS palliation to 53.4±6.6% at stage II in a larger population of HLHS patients (43), a trend attributable to the changes in loading conditions imposed by the stage II operation. Notably, global longitudinal strain (GLS), a load independent marker of myocardial contractility, remained unchanged throughout the follow up period compared to baseline. This is a promising observation since a lower GLS is associated with death and heart transplantation in HLHS patients (44). A significant reduction in CMR-derived tricuspid regurgitant fraction was observed at 6- and 12-months follow-up. We also observed a trend toward decreasing BNP levels in the year following BDCPA. BNP has been well demonstrated as a marker of cardiac dysfunction in patients with single-ventricle physiology (45, 46), so this trend may suggest a progression toward improved cardiac function.

To gain molecular correlates of the observations, we measured RNA content of circulating exosomes in a sub-group of patients, and used computational modeling not only to determine whether 6-to-12 month functional changes can be predicted from early biomarkers, but also to understand potential underlying mechanisms. Interestingly, samples taken a short time after cell delivery (within a week) were able to model long-term functional changes: we constructed a multivariate regression model and identified clusters of co-expressed genes correlated to outcomes. While some computational tools can overfit data and force relationships, we used both guided and unsupervised learning methods to explore these data and identified exosomal RNAs deemed ‘important’ in both analyses. To our understanding, this is the first report of isolating cell therapy-derived exosomes from treated children and of using the data to determine important early biomarkers. We previously published this methodology using human cells in a rat model, and now have extended it to human clinical trials (47). The ability to make predictions early in the therapeutic timeline could allow clinicians to noninvasively monitor potential improvements and, perhaps, even inform ongoing medical treatments. As we are presently conducting a phase II randomized trial of this approach in HLHS, the ability to make a priori predictions could be quite powerful and allow better model refinement and validation.

There are several caveats to our computational approach, including a small sample size (*N*=6) due to the nature of the phase I trial, and the model merely predicts potential mechanisms, and requires additional validation. These mechanisms may not be true functional mechanisms but may instead be function improvement biomarkers. Future studies will need to confirm whether these signals have functional effects markers of cardiac function.

ELPIS limitations include a small sample size and the absence of placebo-treated patients. These are common to many phase I trials, which result from the novel nature of the treatment and difficulty with masking because of the study intervention and design. We emphasize that ELPIS was designed to investigate the intramyocardial MSC infusion safety and feasibility in HLHS patients, and not the efficacy. All efficacy data are hypothesis generating intended to inform the design of larger controlled studies. Furthermore, the importance of physiological changes created by the surgical connection of the superior vena cava to the pulmonary artery on the RV function is unclear because of limited previous serial CMR studies in HLHS patients.

In conclusion, the ELPIS I results suggest that allogeneic MSCs intramyocardial infusion in HLHS patients is feasible, safe, and may have promising effects on the RV through 1-year after treatment. Since ELPIS is the first study of MSCs in HLHS infants, the results are important for advancing this new form of cell therapy for all ages and congenital heart conditions. The data from our study warrant further, appropriately powered phase II studies which are currently underway.

## Data Availability

All data produced in the present study are available upon reasonable request to the authors

## PERSPECTIVES

### COMPETENCEY IN PATIENT CARE AND PROCEDURAL OUTCOME

The ELPIS Phase I study demonstrates the feasibility and safety of administrating allogenic MSCs to HLHS babies at the time of the second Glenn operation. The study also suggests that exosomes specific to the transplanted MSCs can be detected in the serum of treated babies which provides a liquid biopsy for possible mechanistic pathways of myocardial remodeling.

### TRANSLATIONAL OUTLOOK

Randomized studies are needed to verify the efficacy of the allogeneic MSCs in HLHS babies.

## ABBREVIATIONS

BDCPA: bidirectional cavopulmonary anastomosis
BNP: brain natriuretic peptide
BSA: body surface area
CMR: cardiac magnetic resonance imaging
Phase 1 ELPIS Trial: Allogeneic Human Mesenchymal Stem Cell Injection in Patients with Hypoplastic Left Heart Syndrome: An Open Label Pilot Study
GLS: global longitudinal strain
HLHS: hypoplastic left heart syndrome
HAS: human serum albumin
HIV: human immunodeficiency virus
MACE: major adverse cardiac events
MSCs: medicinal signaling cells
PCA: principal component analysis
PLSR: partial least squares regression
RNA: ribonucleic acid
RV: right ventricle/right ventricular
RVDd: right ventricular end-diastolic diameter
RVDs: right ventricular end-systolic diameter
RVEDV: right ventricular end diastolic volume
RVEF: right ventricular ejection fraction
RVESV: right ventricular end systolic volume
RVSV: right ventricular stroke volume
TOM: topological overlap matrix
TR: tricuspid regurgitation
TR RF: tricuspid regurgitant fraction
WGCNA: weighted gene co-expression network analysis

---

**Supplemental Table 1.** ELPIS phase I clinical outcomes. (Lomecel-B)

| Measure | Baseline (n=10) | 6-month (n=9) | 12-month (n=8) | 6-month Change from Baseline | 12-month Change from Baseline |
| --- | --- | --- | --- | --- | --- |
| BSA (m<sup>2</sup>) | 0.331±0.03 | 0.432±0.03 | 0.478±0.034 | 0.105±0.03* | 0.144±0.037*† |
| Length for Age Z-Score | -0.9±2.0 | 0.2±1.3 | -1.2±0.8 | 1.24±1.64 | -0.40±1.83‡ |
| Weight for Age Z-Score | -1.1±1.3 | -0.3±1.0 | -0.6±0.8 | 0.83±1.28 | 0.23±1.13 |
| RV Ejection Fraction (%) | 62.62±5.99 | 53.69±9.56 | 52.31±5.63 | -8.89±10.93 | -10.88±10.7 |
| RV End Systolic Volume Index (mL/m<sup>2</sup> BSA<sup>1.3</sup>) | 49.71±15.61 | 69.85±19.87 | 63.03±13.39 | 18.69±24.2 | 15.3±19.42 |
| RV End Diastolic Volume Index (mL/m<sup>2</sup> BSA<sup>1.3</sup>) | 133.59±36.14 | 152.07±32.09 | 133.44±31.42 | 14.65±46.73 | 2.55±48.04 |
| RV Stroke Volume Index (mL/m<sup>2</sup> BSA<sup>1.3</sup>) | 83.88±23.65 | 82.22±25.25 | 70.41±20.5 | -4.05±33.35 | -12.75±34.8 |
| RV Mass Index (g/m<sup>2</sup> BSA<sup>1.3</sup>) | 101.26±33.71 | 110.82±29.63 | 97.88±24.22 | 7.69±30.22 | 2.23±33.48 |
| TR fraction | 0.45±0.19 | 0.32±0.06 | 0.06±0.09 | -0.09±0.33 | -0.52±0.07 |
| RV systolic diameter (mm/m<sup>2</sup> √BSA) | 37.5±9.24 | 45.76±11.79 | 41.54±8.59 | 7.75±8.89 | 3.84±7.98 |
| RV diastolic diameter (mm/m<sup>2</sup> √BSA) | 61.36±13.95 | 64.48±11.03 | 62.61±9.44 | 2.17±8.44 | -0.65±8.41 |
| GLS (%) | -24.39±6.99 | -20.55±3.05 | -23.88±4.6 | 2.6±8.25 | 0.06±10.76 |
| Sphericity Index | 1.31±0.35 | 1.21±0.26 | 1.22±0.21 | -0.1±0.17 | -0.02±0.19 |

BSA: body surface area; GLS: global longitudinal strain; RV: right ventricle \*p<0.0001 vs baseline, and †p<0.01, ‡p<0.05 vs 6-months using One-Way ANOVA with Mixed-Effects Model for multiple comparisons and Bonferroni correction.

**Supplemental Table 2.** ELPIS phase I clinical outcomes. (Allo-MSCs)

| Measure | Baseline (n=4) | 6-month (n=4) | 12-month (n=4) | 6-month Change from Baseline | 12-month Change from Baseline |
| --- | --- | --- | --- | --- | --- |
| BSA (m²) | 0.327±0.032 | 0.415±0.038 | 0.458±0.040 | 0.088±0.023 * | 0.131±0.028† |
| Length for Age Z-Score | -1.350±0.719 | -1.400±1.745 | -0.450±1.462 | -0.050±1.173 | 0.900±1.042 |
| Weight for Age Z-Score | -0.450±0.656 | 0.125±0.263 | -0.600±1.030 | 0.575±0.629 | -0.150±0.714 |
| RV Ejection Fraction (%) | 67.97±11.02 | 58.36±10.81 | 56.46±16.55 | -9.77±3.46 | -11.52±9.09 |
| RV End Systolic Volume Index (mL/m² BSA<sup>1.3</sup>) | 61.38±48.43 | 91.19±63.78 | 89.95±77.72 | 26.65±5.60* | 28.57±36.06 |
| RV End Diastolic Volume Index (mL/m² BSA<sup>1.3</sup>) | 170.25±85.19 | 204.53±100.14 | 184.38±81.51 | 30.75±20.68 | 14.13±40.91 |
| RV Stroke Volume Index (mL/m² BSA<sup>1.3</sup>) | 108.87±37.03 | 113.34±37.36 | 94.44±8.91 | 4.10±19.96 | -14.44±38.23 |
| RV Mass Index (g/m² BSA<sup>1.3</sup>) | 116.15±34.97 | 133.54±39.34 | 105.40±45.95 | 7.94±4.12 | -10.75±16.60 |
| TR fraction<sup>a</sup> | 0.51±0.20 | 0.36±0.37 | 0.32±0.20 | -0.15±0.17 | -0.19±0.14 |
| RV systolic diameter (mm/m² √BSA) | 40.49±22.07 | 52.60±28.66 | 49.66±20.24 | 9.40±6.97 | 9.17±7.29 |
| RV diastolic diameter (mm/m² √BSA) | 68.00±21.56 | 76.61±21.48 | 68.78±21.47 | 4.68±3.47 | 0.78±7.58 |
| GLS (%) | -21.29±6.65 | -20.91±3.36 | -24.30±3.75 | -2.93±3.12 | -3.01±2.90 |
| Sphericity Index | 1.30±0.26 | 1.05±0.21 | 1.38±0.32 | -0.19±0.30 | 0.08±0.16 |

BSA: body surface area; GLS: global longitudinal strain; RV: right ventricle \*p<0.05, †p<0.01vs baseline using One-Way ANOVA with Mixed-Effects Model for multiple comparisons and Bonferroni correction.

